# Dietary Factors Affect Brain Iron Accumulation and Parkinson’s Disease Risk

**DOI:** 10.1101/2024.03.13.24304253

**Authors:** Jonathan Ahern, Mary ET Boyle, Leo Sugrue, Ole Andreassen, Anders Dale, Wesley K. Thompson, Chun Chieh Fan, Robert Loughnan

## Abstract

**Background:** Alterations in brain iron accumulation, representing both excess or insufficiency of iron, are associated with Parkinson’s disease. Iron is exclusively absorbed from food; therefore, dietary patterns may impact risk.

**Objective:** Assess associations between dietary preferences, brain iron, and Parkinson’s disease risk.

**Methods:** This study included 228,653 individuals and utilized dietary surveys, magnetic resonance imaging, and electronic health records of neurological diagnoses. We performed regression analyses using i) estimated dietary nutrient intake, ii) dietary preferences, and iii) a lower-dimensional space factor-loading dietary preferences to predict a novel brain iron accumulation score derived from T2-weighted MRI and Parkinson’s disease diagnosis.

**Results:** We found strong positive associations between measures of brain iron accumulation and the alcohol factor (*t*=4.02, *p_FDR_*<0.001) and significant negative associations between brain iron accumulation and the high-sugar factor (*t*=-3.73, *p_FDR_*<0.001). Conversely, the alcohol factor was associated with decreased Parkinson’s disease risk (*t*=-5.83, *p* <1×10^−8^) and the high-sugar factor was associated with increased Parkinson’s disease risk (*t*=6.03, *p* <1×10^−8^). Instrumental variable regression revealed that dietary preferences associated with higher brain iron were also associated with lower risk for Parkinson’s disease (□=-0.247, *p*=0.004). Voxel-wise imaging analysis displayed iron accumulation patterns consistent with these findings.

**Conclusion:** Contrary to some prior studies, we find lower central iron levels to be associated with greater Parkinson’s disease risk. Dietary components, especially alcohol and carbohydrates, are related to differences in brain iron accumulation in motor regions of the brain and differences in risk of Parkinson’s disease, suggesting the potential for dietary interventions to influence Parkinson’s disease risk.

Graphical Abstract:
Dietary and Lifestyle Factors of Brain Iron Accumulation *Previous research has established that iron overload in the brain is associated with increased risk of movement disorders.^4–7^

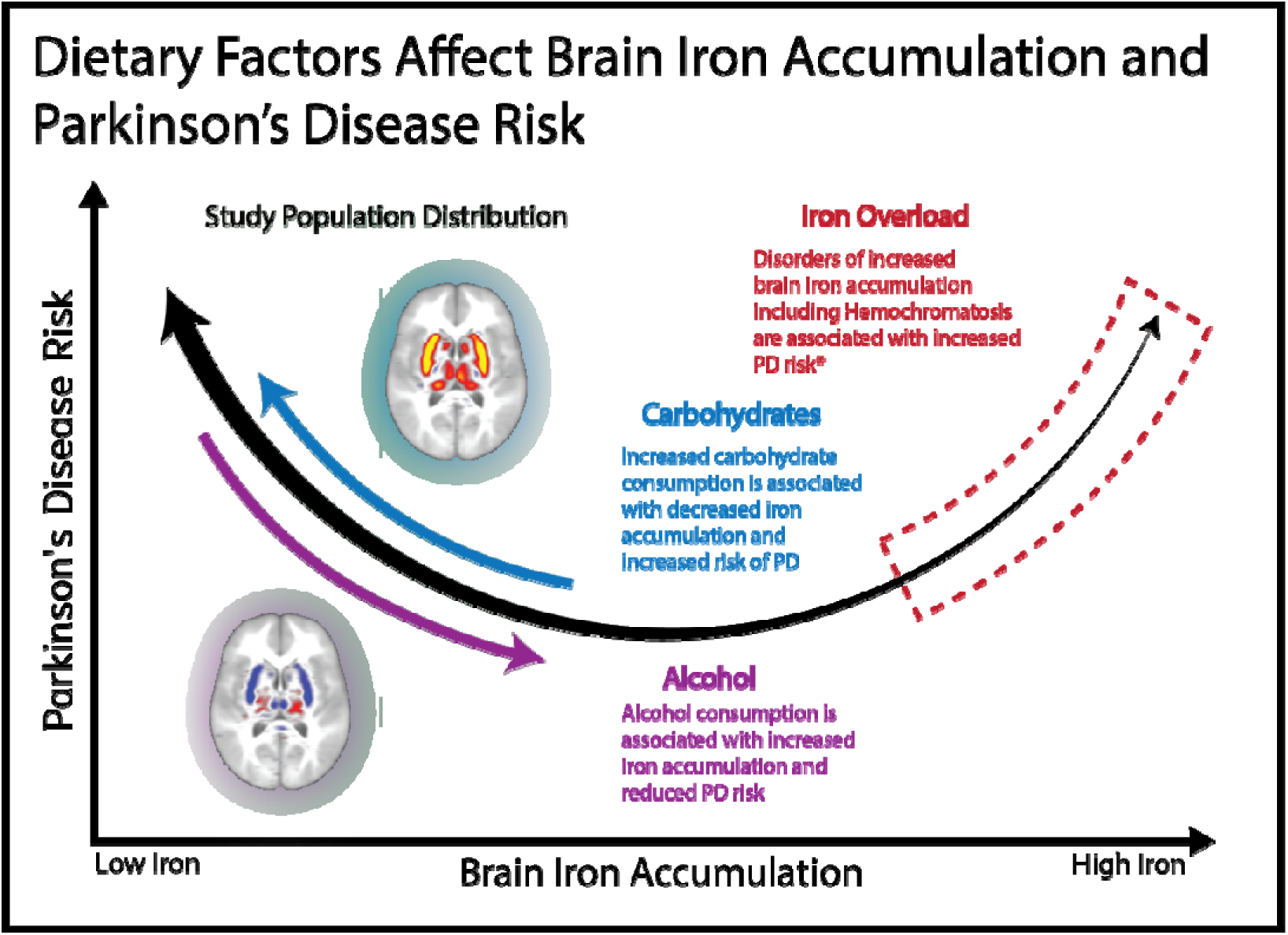

## Introduction

Parkinson’s disease (PD) is the fastest-growing neurodegenerative disease^1^ and is characterized by progressive movement difficulties^2^ and reduction in quality of life.^3^ Iron dyshomeostasis is associated with movement disorders,^4–7^ with increased iron accumulation in regions like the substantia nigra and the basal ganglia being common findings in PD patients.^8,9^ While iron is an essential nutrient, iron overload can lead to ferroptotic mechanisms implicated in PD.^10,11^

Low peripheral iron is associated with greater PD risk.^11–14^ Conversely, hemochromatosis, characterized by iron overload,^15^ has also been linked to an increased PD risk.^4,6^ These studies suggest a U-shaped relationship between iron and PD risk, with both low and high levels being linked to increased risk. A recent study using neuroimaging in a large cohort^6^, found that lower central iron levels were linked to a higher PD risk in individuals without hemochromatosis. Conversely, higher central iron levels were associated with increased PD risk in individuals carrying hemochromatosis risk alleles. Under this U-shaped model, a normative cohort, free from major iron-disruption, lies on the descending arm, exhibiting a linear negative central iron–PD risk association (Graphical Abstract).

PD results from an interaction between genetic and environmental factors that accumulate throughout life,^2^ with diet representing an important modifiable factor. Dietary patterns, like the Mediterranean diet,^19^ have been shown to improve PD outcomes.^20,21^ Conversely, extended consumption of ultra-processed foods is related to increased risk of nonmotor features in PD.^22^

We aim to investigate whether diet impacts PD risk through changes in brain iron. We build upon work identifying a brain iron-specific biomarker of PD referred to as the Hemochromatosis Brain PolyVoxel Score (HB-PVS). This marker quantifies the central iron dysregulation associated with hemochromatosis, as captured by T2-weighted MRI (T2w-MRI), in motor regions of the brain.^6,7^ The HB-PVS is differentially associated with PD risk in individuals with and without hemochromatosis. Individuals with hemochromatosis and very high HB-PVS show increased PD risk, whereas in individuals without hemochromatosis, a lower HB-PVS compared to other subjects is associated with increased PD risk.^6^

## Materials and methods

### UK Biobank Measures

Genotypes, neuroimaging, demographics, diagnoses, and dietary measures were obtained from the UK Biobank (URLs) under accession number 27412, excluding 206 participants who withdrew their consent. All participants provided electronically signed informed consent. The study was approved by the UK Biobank Ethics and Governance Council. The recruitment period for participants was from 2006 to 2010, and participants had to be 40-69 years old during this period to be included in the sample.

### Neuroimaging Data

#### Image Processing

Image acquisition and processing are described in the supplementary methods. The average of the *b*=0 images from registered diffusion-weighted MRI intensities (dw-MRI) was used as voxel-wise measures of T2w-MRI. T2w-MRI refers to the average of *b*=0 images, and dw-MRI refers to the full diffusion acquisition (all b directions and values). Using a restriction spectrum imaging (RSI)^23^ model, we separated dw-MRI signal into three compartments: intracellular, extracellular, and unhindered free water. This model estimates fractions of diffusion attributable to these compartments using a mixture of spherical harmonic basis functions.^24^ Although prior work indicated that the HB-PVS, derived from T2w-MRI, is sensitive to iron,^6,7^ T2w-MRI signal can also be sensitive to other cellular changes, such as gliosis and edema,^25^ pathologic processes that may reflect ischemic injury or inflammation.^26^ We used the free water RSI component in sensitivity analysis to distinguish iron-related from non-iron, vascular-related signals for discovered associations.

#### The Hemochromatosis Brain PolyVoxel Score

The primary imaging measure is the HB-PVS, derived from a classifier trained on UK Biobank T2w-MRI brain scans, which are sensitive to iron,^27^ to differentiate between individuals with and without C282Y homozygosity.^6^ C282Y homozygosity accounts for 95% of hemochromatosis cases,^28^ a disorder that can lead to iron overload.^15^ As detailed by Loughnan *et al.*,^6^ the HB-PVS quantifies iron accumulation across motor-related brain regions, including the cerebellum, thalamus, caudate, putamen, pallidum, red nucleus, substantia nigra, and subthalamic nucleus (Figure S1).

### Preference Data

Preference data (Category 1039) were gathered from a web-based questionnaire of preferences for foods and activities. The questionnaire included 150 items measured on a nine-point scale from ‘extreme dislike’ to ‘extreme like’.^29^ After noninformative values (‘Never tried’ or ‘Prefer not to answer’) were removed, six preferences were removed for having greater than 10% missingness across the population (preferences for bell peppers, capers, globe artichoke, going to the gym, pollock, and soy milk), and 3,459 participants were removed for having individual missingness of greater than 10%. Remaining missing values were imputed from the median value for that variable across the remaining subjects. Data were z-score normalized across all remaining subjects.

### Nutrient Data

As a confirmatory analysis, we analyzed the effects of estimated nutrient intake (Category 100117; estimated independently from preference data) on the HB-PVS and PD risk. Further details can be found in the supplementary methods.

### Disease Diagnosis

Neurological diagnoses were extracted from ICD-10 diagnosis data from electronic health records (Data-Field 41270). PD diagnosis included diagnosis codes: F02.3, G20, G21.0, G21.1, G21.2, G21.3, G21.4, G21.8, and G21.9. Vascular Dementia (VD) diagnoses, which were used as a positive control in our voxel-wise imaging analysis, included the diagnosis codes: F01.0, F01.1, F01.2, F01.3, F01.8, and F01.9. Individuals with one or more of these diagnoses were categorized as having been diagnosed with PD (Table S2) or VD, respectively.

### Statistical Analyses

Analyses were performed using generalized linear regressions in Python using statsmodels (version 0.14.0). All analyses were controlled for sex (Data-Field 31), the maximum reported pre-tax income category (Data-Field 738), and the top ten genetic principal components (Data-Field 22009). Analyses that involve the HB-PVS additionally controlled for scanner site (Data-Field 54) to account for differences in imaging equipment. All analyses controlled for age; imaging analyses controlled for the age during the scan (Data-Field 20216), and disease analyses of preferences and nutrition were corrected for age during survey completion (Data-Field 20750 and 105010). *P*-values within each analysis were corrected using the multitest correction from statsmodels using Benjamini/Hochberg False Discovery Rate (FDR) and Bonferroni (Bonf.) correction methods.

Lower dimensional factors of preference data were generated in Python using Educational Testing Service’s Factor_Analyzer package (version 0.5.0) with default parameters, which calculates factors by minimizing residuals^30,31^. Factors were varimax rotated to increase interpretability^32^. To calculate robust standard errors, we performed 10,000 bootstrap iterations of our regression analyses^33^.

To test whether the effect of dietary preferences on PD risk was mediated by brain iron concentration, we performed two-sample instrumental variable mediation analysis^34,35^. In this framework, individual preferences are used as instruments to test the role of mediating traits on outcomes. Here, we use preference questionnaire data as instrumental exposures, PD risk as outcome, and brain iron concentration (from the HB-PVS) as the mediator to test the regression:

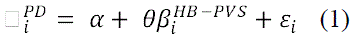

Where 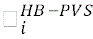 and 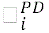 are the coefficients associating the *i^th^* food preference question with the HB-PVS and PD diagnosis, respectively, calculated from previous analyses. α is the intercept, □*_i_* is the error, and □ is the mediation effect. We interpret the results as suggestive of dietary factors impacting PD risk through brain iron levels, but refrain from making strong claims about causality.

Our analysis is focused on dietary factors, so we removed eight preference questions related to non-dietary factors (preferences for taking the stairs, working up a sweat, exercising alone, bicycling, going to a cafe, watching television, cigarette smoking, exercising with others, and going to the pub) — some of which (exercise in particular) we expected to be susceptible to reverse causality with PD risk. An analysis including non-dietary questions is included in Figure S8.

Statistical analysis for our confirmatory nutrient analysis is largely consistent with the methods described above (supplementary methods).

### Voxel-wise Associations of Dietary Factors

We generated voxel-wise association maps between T2w-MRI intensities^27^ and dietary preference factors two (sweets) and seven (alcohol). To account for the potential microvascular pathologies, which we expect to be preferentially related to differences in the free water fraction of RSI, we generated association maps between normalized free water diffusion from RSI models and these dietary factors.

Association maps were generated by voxel-wise univariate regression using all 28,388 individuals from our preference imaging subsample (Table 1), predicting T2w-MRI and free water signal from dietary factors two (sweets) and seven (alcohol), separately. Covariates of age at scan, sex, scanner, and the top ten principal components of genetic ancestry were corrected by pre-residualizing images before fitting. We plotted voxel-wise effect sizes (□) from these models after passing a smoothing kernel with a full width at half maximum of 2.82 voxels. Effect size values were masked to those used to compute the HB-PVS.

**Table 1:**
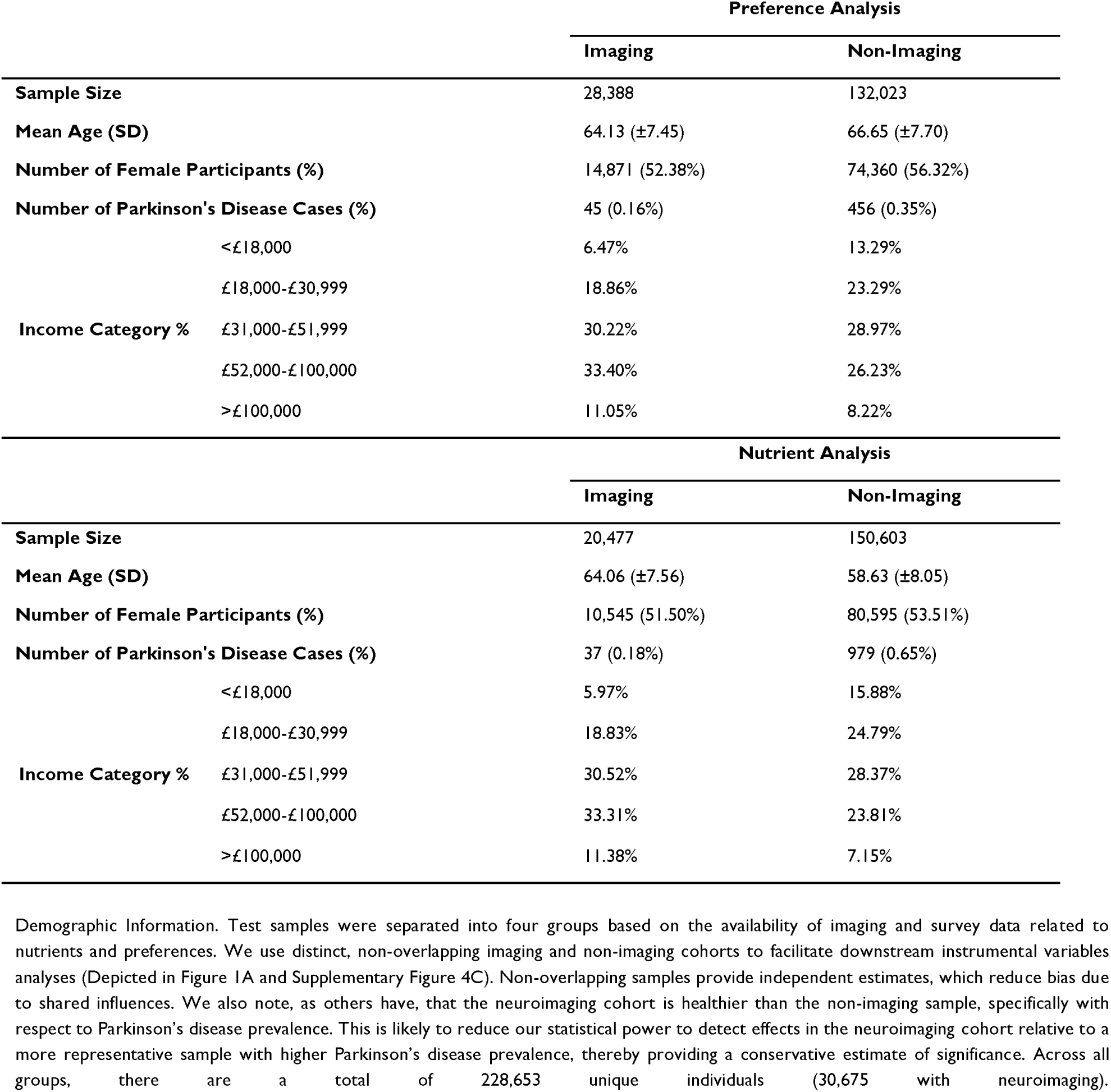
Demographics of Imaging and Non-Imaging Cohorts.

## Results

A total of 228,653 individuals were included in this study across four cohorts (Table 1).

### Preference Analysis

We fit a series of linear and logistic regression models associating each preference with the HB-PVS and PD risk. Here, we describe general trends among our preference findings. Full results can be found in the Supplementary Results (Figures S2-S3, Tables S5-S6).

Increased HB-PVS was associated with preferences for alcohol (red wine: *t*=6.72, *p_FDR_*<1×10^−8^; white wine: *t*=5.48, *p_FDR_*<1×10^−5^) and red meats (red meat: *t*=4.46, *p_FDR_*<1×10^−3^; lamb: *t*=4.30, *p_FDR_*<1×10^−3^). Decreased HB-PVS was associated with preferences for cereal grains (corn flakes: *t*=-4.61, *p_FDR_*<1×10^−5^; porridge *t*=-3.74, *p_FDR_*=0.004), sweet foods (biscuits: *t*=-3.64, *p_FDR_*=0.004; cake: *t*=-3.57, *p_FDR_*=0.004), and exercise (bicycling: *t*=-3.19, *p_FDR_*=0.010; exercising alone: *t*=-3.03, *p_FDR_*=0.015).

Decreased PD risk was associated with exercise preferences (taking the stairs: *t*=-10.87, *p_FDR_*<1×10^−24^; working up a sweat: *t*=-7.67, *p_FDR_*<1×10^−10^), vegetables (salad leaves: *t*=-8.27, *p_FDR_*<1×10^−13^; vegetables: *t*=-6.24, *p_FDR_*<1×10^−7^), and alcoholic beverages (spirits: *t*=-6.15, *p_FDR_*<1×10^−7^; red wine: *t*=-5.80, *p_FDR_*<1×10^−6^). Increased PD risk was associated with preferences for sweet foods (sweet coffee house drinks: *t*=5.15, *p_FDR_*<1×10^−5^; cake icing: *t*=4.90, *p_FDR_*<1×10^−4^).

### Dietary Preference Effects on Brain Iron Predict Parkinson’s Risk

We use the HB-PVS as a biomarker of brain iron levels. Details of its development and training are provided elsewhere.^6^ In brief, contributions to the HB-PVS are determined by region-specific classifier weightings (Figure S1). Higher HB-PVS scores generally reflect higher iron concentrations across most brain regions. However, certain regions that contribute less to the score, namely the caudate and substantia nigra, may exhibit lower iron concentrations at higher HB-PVS values. Thus, the HB-PVS may also capture abnormal patterns of iron distribution within the brain. Overall, previous work has shown that the HB-PVS correlates strongly with genes involved in iron metabolism, peripheral iron levels, and PD.^6,7^

We hypothesized that the effect of specific foods on PD risk was mediated through their impact on brain iron concentration. We performed mediation analysis, using two-sample instrumental variable estimation. Here, preferences were regarded as exposures, PD as the outcome, and brain iron accumulation (HB-PVS) as the mediator. When restricting to dietary preferences, coefficients from HB-PVS and PD display a significant negative association (Figure 1A: □=-0.247, *p*=0.004), indicating that preferences associated with higher brain□iron accumulation tended to confer lower PD risk, and vice versa. This negative β - β relationship is consistent with a model in which dietary modulation of central iron contributes to PD susceptibility. We emphasize, however, that these results are exploratory; while the strength and direction of the association support an iron□mediated pathway (diet to brain iron to PD risk), residual direct effects of diet on PD risk and potential confounders cannot be fully excluded without further validation. This trend does not persist when including non-dietary preferences in our analysis for reasons we believe to be related to reverse causation (Supplementary Results and Figure S8).

**Figure 1:**
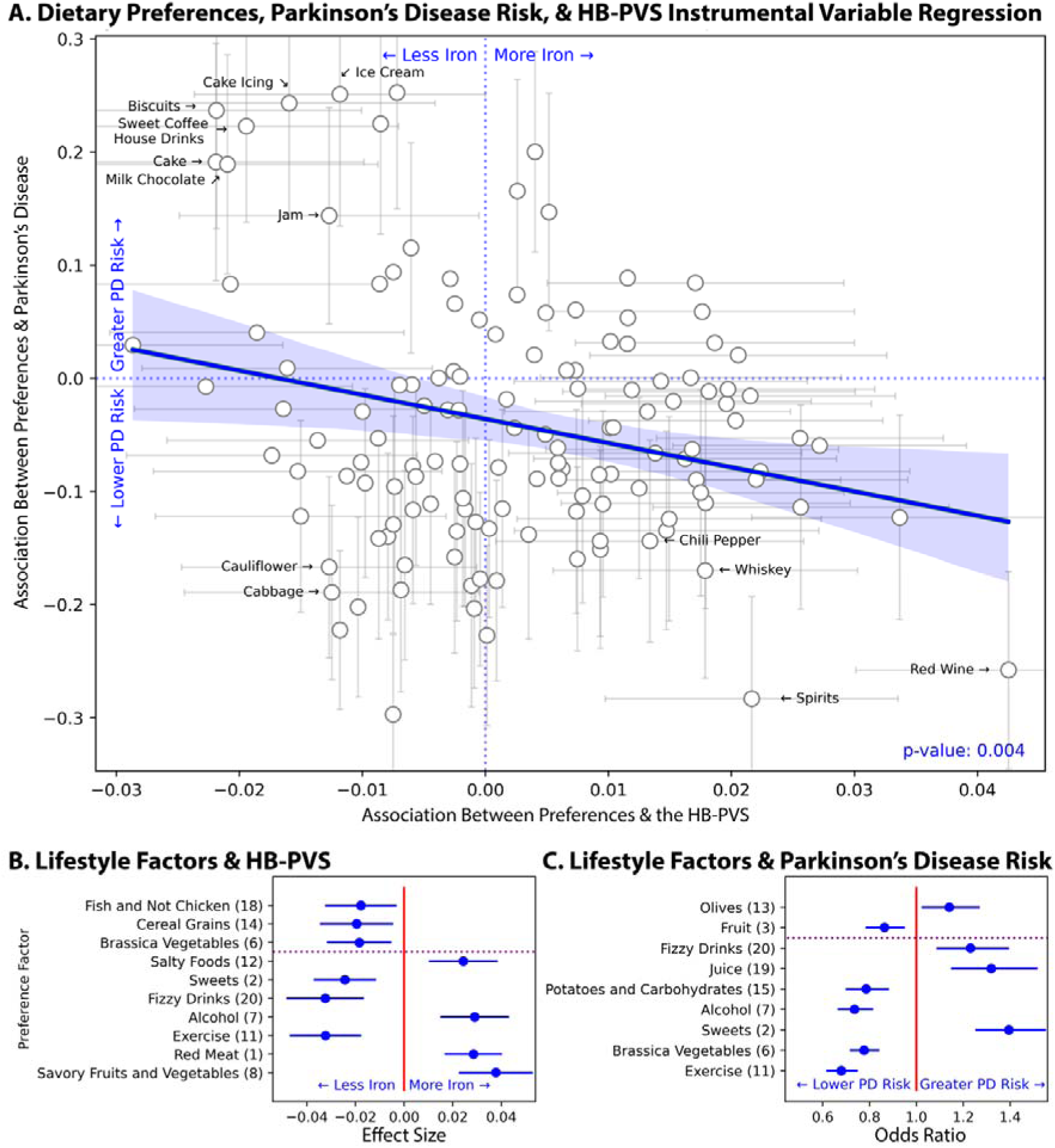
Preference Analysis. (A) Scatter plot of the instrumental variable regression (i) between dietary preferences and the HB-PVS and (ii) preferences and the odds ratio of developing PD. Error bars are shown for factors that are significant along a given axis. Of those factors, the top thirteen dietary preference factors furthest from the origin are annotated. A regression line shows the correlation between the effects of a dietary preference on PD and the HB-PVS, respectively. A scatter plot of the dietary and non-dietary factors can be found in Figure S8. (B) Forest plot showing the FDR associations between preference factors and the HB-PVS. Results below the dotted line are additionally Bonferroni Significant. A full numeric summary of the effects of all factors can be found in Table S8. (C) Forest plot showing the FDR associations between preference factors and odds ratio of developing PD. Results below the dotted line are additionally Bonferroni Significant. A full numeric summary of the effects of all factors can be found in Table S9.

### Nutrient Analysis

Analysis of estimated nutrient intake supported several findings from the preference analysis. We found that estimated intakes related to sugars were associated with decreased HB-PVS (maltose: *t*=-3.74, *p_FDR_*=0.012; lactose: *t*=-3.36, *p_FDR_*=0.013) and increased PD risk (carbohydrates: *t*=3.46, *p_FDR_*=0.012; total sugar: *t*=3.45, *p_FDR_*=0.012). Estimates related to alcohol were associated with decreased PD risk (*t*=-4.56, *p_FDR_*<0.001), in line with preference findings. Instrumental variable analysis showed a general downward trend (□=-0.421, *p*=0.0006) where nutrient intake associated with decreased HB-PVS was associated with increased PD risk (Supplementary Results, Figure S4, and Tables S3-S4).

### Factor Analysis of Preferences

Due to the high correlation structure in dietary preferences (Figure S5), we performed factor analysis to identify interpretable latent factors. Twenty factors accounted for approximately half (49.77%) of the variance in the preference space (Table 2, Figure S6). For full loadings, see Figure S7 and Table S7.

**Table 2:** Preference Factor Loadings.

| **Factor Number** | **Top Preferences (Loading)** | **Interpretation** | **Variance Explained** | **Cumulative Variance Explained** |
| --- | --- | --- | --- | --- |
| **Factor 1** | red meat (0.844), beef steak (0.790), barbequed or grilled meat (0.765) | Red Meat | 11.77% | 11.77% |
| **Factor 2** | cake (0.794), sweet foods (0.756), biscuits (0.709) | Sweets | 8.20% | 19.97% |
| **Factor 3** | fruit (0.717), plums (0.672), oranges (0.577) | Fruit | 4.43% | 24.40% |
| **Factor 4** | haddock (0.743), baked steamed fish (0.735), mackerel (0.721) | Fish | 4.11% | 28.51% |
| **Factor 5** | spicy foods (0.886), chili pepper (0.790), burn of spicy foods (0.746) | Spicy Foods | 2.30% | 30.81% |
| **Factor 6** | cabbage (0.711), cauliflower (0.685), Brussel sprouts (0.641) | Brassica Vegetables | 2.00% | 32.81% |
| **Factor 7** | spirits (0.648), whisky (0.604), red wine (0.578) | Alcohol | 1.88% | 34.68% |
| **Factor 8** | avocados (0.509), aubergine (0.465), asparagus (0.443) | Savory Fruits & Vegetables | 1.74% | 36.42% |
| **Factor 9** | hard cheese (0.596), soft cheese (0.589), blue cheese (0.468) | Cheese | 1.54% | 37.96% |
| **Factor 10** | tea with sugar (-0.732), coffee with sugar (-0.731), coffee without sugar (0.541) | Unsweetened Tea & Coffee | 1.42% | 39.38% |
| **Factor 11** | working up a sweat (0.753), exercising alone (0.529), bicycling (0.497) | Exercise | 1.30% | 40.68% |
| **Factor 12** | salty foods (0.701), adding salt to foods (0.526), salty pretzels (0.463) | Salty Foods | 1.24% | 41.93% |
| **Factor 13** | green olives (0.767), black olives (0.753), gherkins (0.340) | Olives | 1.23% | 43.15% |
| **Factor 14** | whole grain breakfast cereal (0.481), brown rice (0.351), porridge (0.350) | Cereal Grains | 1.14% | 44.30% |
| **Factor 15** | potatoes (0.377), chips/French fries (0.370), pasta (0.366) | Potatoes and Carbs | 1.04% | 45.34% |
| **Factor 16** | salad dressing (0.504), mayonnaise (0.419), vinegar (0.370) | Condiments | 0.99% | 46.33% |
| **Factor 17** | whole milk (0.496), cream (0.416), dairy products (0.384) | Dairy | 0.93% | 47.27% |
| **Factor 18** | chicken (-0.337), roast chicken (-0.319), herring (0.294) | Fish and Not Chicken | 0.87% | 48.14% |
| **Factor 19** | orange juice (0.501), apple juice (0.399), grapefruit (0.253) | Juices | 0.85% | 48.99% |
| **Factor 20** | diet fizzy drinks (0.356), corn flakes (0.222), regular nondiet fizzy drinks (0.204) | Fizzy Drinks | 0.78% | 49.77% |

#### Preference Factors and Brain Iron

We performed linear regressions in our imaging sample (*N*=28,388) predicting the HB-PVS from the first 20 preference factors. All of the results from the preference-level analysis were recapitulated, namely the positive association between alcohol (factor seven: *t*=4.02, *p_FDR_*<0.001) and red meat (factor one: *t*=4.73, *p_FDR_*<1×10^−4^) and the negative associations for cereal grains (factor fourteen: *t*=-2.54, *p_FDR_*=0.012), sweets (factor two: *t*=-3.73, *p_FDR_*<0.001), and exercise (factor eleven: *t*=-4.31, *p_FDR_*<0.001).

We found additional positive associations between the HB-PVS and savory fruit and vegetables (factor eight: *t*=4.86, *p_FDR_*<1×10^−4^) and salty foods (factor twelve: *t*=3.36, *p_FDR_*=0.001) and negative associations between the HB-PVS and fizzy drinks (factor twenty: *t*=-4.00, *p_FDR_*<0.001), brassica vegetables (factor six: *t*=-2.74, *p_FDR_*=0.008), and preference for fish and a dislike for chicken (factor eighteen: *t*=-2.36, *p_FDR_*=0.018) (Figure 1B, Table S8).

#### Preference Factors and Parkinson’s Disease Risk

Using the same top twenty preference factors, we fit models to predict PD risk in our non-imaging sample (*N_Case_*=456, *N_Control_*=131,567). We recapitulated the results from the preference-level analysis, namely increased PD risk was associated with a lower preference for exercising (factor eleven: *t*=-7.66, *p_FDR_*<1×10^−12^), alcohol (factor seven: *t*=-5.82, *p_FDR_*<1×10^−7^), vegetables (factor six: *t*=-6.11, *p_FDR_*<1×10^−8^), and fruits (factor three: *t*=-2.98, *p_FDR_*=0.018), and higher preference for sweets (factor two: *t*=6.03, *p_FDR_*<1×10^−8^).

We also found that increased PD risk was associated with a lower preference for potato products and carbohydrates (factor fifteen: *t*=-4.03, *p_FDR_*<0.001) and a higher preference for juices (factor nineteen: *t*=3.89, *p_FDR_*<0.001), fizzy drinks (factor twenty: *t*=3.23, *p_FDR_*=0.002), and olives (factor thirteen: *t*=2.35, *p_FDR_*=0.018) (Figure 1C, Table S9).

#### Voxel-wise Association Maps of Sweet and Alcohol Factors

We generated voxel-wise association maps between T2w-MRI and dietary factors two (sweets) and seven (alcohol), which were selected because they were significantly associated with both PD risk and the HB-PVS.

Consistent with earlier results of factor two (sweets) being associated with decreased HB-PVS, voxel-wise analysis demonstrated that this factor was associated with T2w-MRI hyperintensities in movement-related regions, including basal ganglia and cerebellar regions, indicating decreased iron accumulation (Figure 2). Conversely, factor seven (alcohol) was associated with T2w-MRI hypointensities in movement-related regions, including the putamen and cerebellum (Figure 2), consistent with earlier results. Interestingly, we also observe regions of modest hyperintensities in thalamic regions.

**Figure 2:**
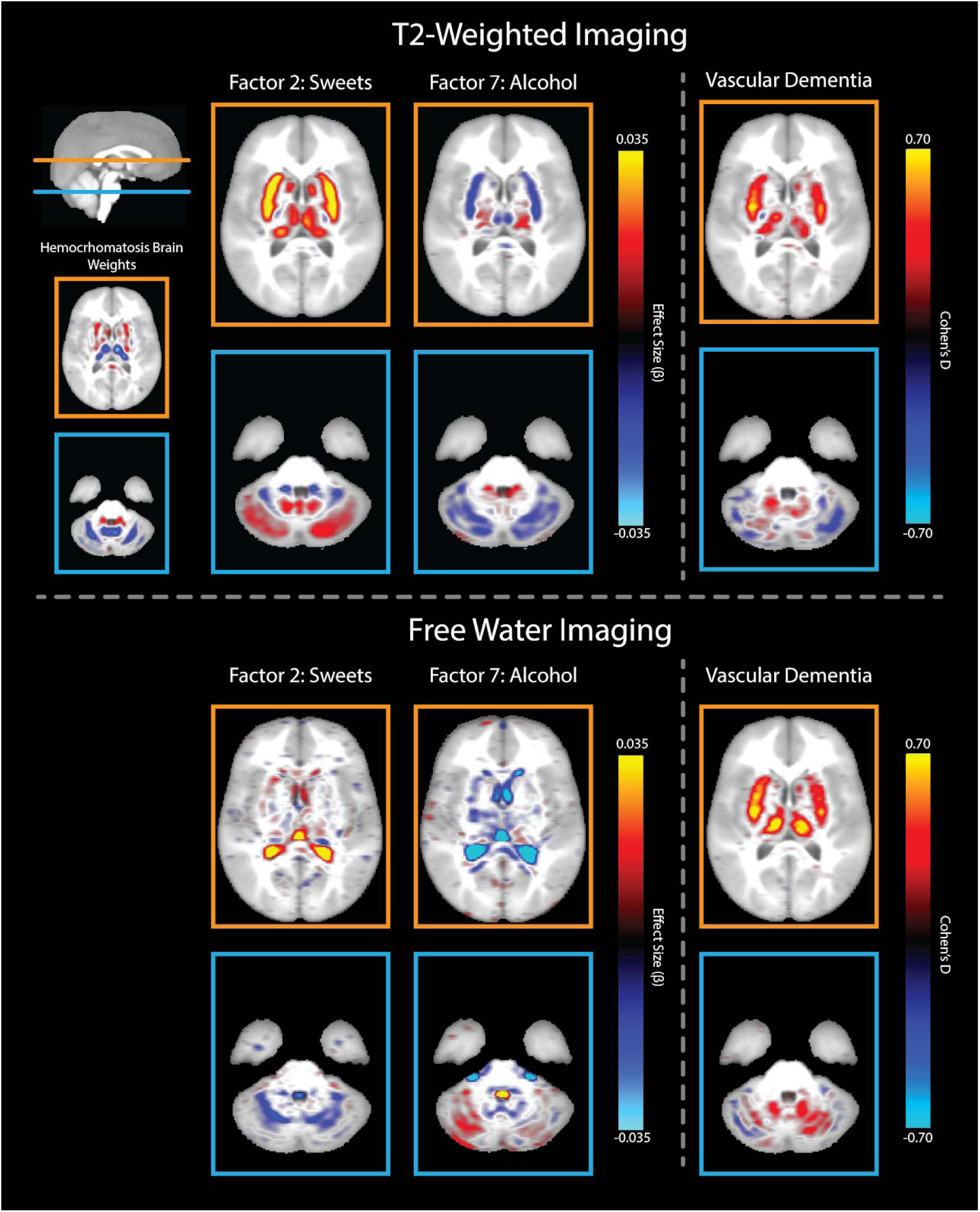
Dietary Factor Association Maps. Brain Voxel-wise association map of T2w-MRI and free water signal intensity for (i) factor two (sweets), (ii) factor seven (alcohol), and (iii) VD as a positive control. Voxels are masked to those that have non-zero weights for HB-PVS (Figure S1). Unmasked factor association maps can be found in Figure S9. The hemochromatosis brain weights (top, far left panels) are included for reference and indicate the relative weighting that intensities in different regions of the brain contribute to the calculation of the HB-PVS, with red regions representing positive weightings and blue regions representing negative weightings, expressed in arbitrary units (Figure S1). In our T2w-MRI brain association maps, effect sizes are displayed. Orange regions represent increased T2w-MRI intensities, consistent with lower iron concentrations for individuals with a higher score along the spectrum of a given dietary factor, and blue regions represent decreased T2w-MRI intensities, consistent with higher iron concentrations. In the free water association maps, higher values (orange) indicate increased isotropic diffusion. The similar association patterns in both T2w-MRI and free water association maps in the VD analysis suggest that changes in free water are likely driving the observed changes in T2w-MRI signal intensity. However, the association maps of the dietary factors show distinct intensity patterns: T2w-MRI signal changes in the basal ganglia and cerebellum that are not associated with changes in free water, suggesting that our T2w-MRI signal changes related to dietary factors are more likely to be driven by iron accumulation differences than vascular differences.

To inspect the degree to which our T2w-MRI associations were driven by vascular pathologies, we associated factors two (sweets) and seven (alcohol) with voxel-wise estimates of free water. Specifically, we were concerned that a high-sugar diet may be related to microvascular pathology.^36,37^ As a positive control, we associated voxel-wise estimates of free water with a VD diagnosis.^38^ We posited that vascular disruptions would increase both T2w-MRI and free water signal, whereas iron differences would only affect the T2w-MRI signal.

Preference factors for sweets and alcohol were associated with basal ganglia and cerebellar intensities for T2w-MRI imaging but not free water estimates (compare the upper and lower panels of Figure 2). For VD, we see higher signal intensities for both T2w-MRI and free water signals (right side of Figure 2). Previous evidence of the HB-PVS’s strong coupling to iron regulation^6,7^ along with these results, indicates that the alcohol and sweet imaging effects are more likely related to iron processes than being confounded by vascular disruptions. Incidental free water hyperintensities are cautiously interpreted in the supplementary results.

## Discussion

Dietary preferences linked to lower brain iron are associated with increased PD risk, aligning with our previous work.^6^ Although this may appear to contradict studies reporting a positive association between brain iron and PD risk^8–10^; our findings offer a broader perspective rather than a conflicting one. While a lower HB-PVS score, reflecting overall reduced brain iron accumulation, is associated with greater PD risk, this does not imply that iron is uniformly reduced across all brain regions. During PVS training, regularization results in some regions contributing negatively and others contributing positively to the overall score (Figure S1). Given these HB-PVS weightings, our results are consistent with higher iron accumulation in regions such as the substantia nigra, pallidum, and caudate being associated with greater PD risk. Simultaneously, larger regions with greater contributions to the HB-PVS, such as the thalamus and cerebellum, show a pattern where lower iron levels are associated with greater PD risk. These regionally specific contributions help reconcile our findings with the broader literature and support a nuanced interpretation of brain iron’s role in PD.

The individual regions that make up the HB-PVS show results consistent with a misdistribution of iron, where in some regions, excessive accumulation is present, and in other larger regions, there is a relative dearth of iron. However, when these results are aggregated, the HB-PVS is linked with lower central iron levels in large areas of the brain and is associated with greater PD risk. The novelty in our approach may capture a more subtle understanding of central iron homeostasis that has been difficult to discover with traditional approaches, allowing us to distill a pathogenic, U-shaped misdistribution of iron into a central tendency that is still sensitive to dietary trends and movement disorders^6^.

### Sugars and Carbohydrates

We found carbohydrate-related preferences were associated with decreases in the HB-PVS, a measure of brain iron, and an increase in PD risk. These findings were supported by our nutrient and factor-level analyses as well as by the T2w-MRI imaging analysis, where we found relative hyperintensity in movement-related regions of the brain, including the putamen and cerebellum, indicating reduced iron accumulation.

Experimental evidence in humans demonstrates a causal relationship between sugar on iron homeostasis, with glucose ingestion decreasing serum iron through pancreatic secretion of the iron-regulatory factor, hepcidin,^39^ consistent with our findings. Epidemiological research shows that glycemic dysregulation and related diseases are related to increased PD risk and worse PD outcomes,^40–42^ in line with our findings of preferences toward sugary foods being related to greater PD risk. Further, iron stores and regulatory molecules contribute to impaired glycemic regulation,^43^ possibly indicating a bidirectional coupling between iron and glucose homeostasis. Taken together, our findings indicate that consumption of sweet and sugary foods contributes to an iron-depleted phenotype in the brain. This iron depletion may, then, contribute to increased PD risk. These results provide a mechanistic interpretation of high-sugar diets as a potential modifiable factor mediating PD risk.

Sugars and carbohydrates may also impact PD risk and iron accumulation at the microbiome level, which is associated with health,^44,45^ including PD.^46,47^ Preferences for sweets and carbohydrates may elevate levels of pro-inflammatory, opportunistic pathogenic bacteria in the gut, which is linked to an increased PD risk.^48^ This dietary pattern also contributes to □-synuclein pathology in the enteric and central nervous system,^49^ where it contributes to ferroptosis, iron-dependent programed cell death, in the brain.^50^

### Alcohol

Preferences related to alcohol were associated with greater brain iron, as evidenced by the HB-PVS, and decreased PD risk. These findings were supported by our nutrient and factor-level analyses. Alcohol-preference-related increases in brain iron in regions associated with the hemochromatosis brain were supported by T2w-MRI imaging, which found hypointensities in the putamen and cerebellum, consistent with higher iron accumulation in those areas associated with greater preference for alcohol. Interestingly, we also found modest hyperintensities in thalamic regions, indicating that alcohol may have differential effects on subregions of the basal ganglia.

There is extensive literature suggesting that moderate alcohol consumption is associated with decreased PD risk,^51,52^ with one meta-analysis suggesting a U-shaped risk profile.^51^

Alcohol consumption is known to impact iron absorption and accumulation. Alcohol can lead to increased iron accumulation in the brain.^53^ Alcohol can downregulate the synthesis of hepcidin, the primary iron-regulating hormone, and, with excessive consumption, can cause iron overload in otherwise typical individuals.^54–57^ In individuals with iron overload disorders, including hemochromatosis, alcohol consumption is associated with worse health outcomes.^58–60^ Conversely, in genetically typical individuals, the moderate consumption of alcohol (fewer than 2 drinks daily) is associated with decreased risk of iron deficiency and its associated anemia.^57^ Anemia is also associated with increased PD risk in some^61–64^ but not all^65^ previous studies.^66^ Measures of iron in the brain have been suggested to have a U-shaped association with PD risk.^6^ Our findings support a more scoping view of the relationship between iron, alcohol, and PD, where alcohol consumption may alter iron-regulation, increasing iron absorption and decreasing anemia, which together may decrease PD risk.

See supplementary materials for a discussion of additional results.

### Limitations

Our dietary measures are self-reported, which is susceptible to reporting bias. Nevertheless, the convergence of results from our preference, nutrient, and factor-level analyses gives us confidence that we are capturing true effects.

The HB-PVS is a univariate measure that is specifically tuned to iron disruptions in specific brain regions that are related to motor control and predictive of PD^27^ risk, including the substantia nigra, thalamus, putamen, and cerebellum^6^. Collapsing the signal across multiple brain regions into a single measure enables the analyses showcased here; however, there is an inherent loss of specificity when performing this dimensionality reduction. The PVS represents larger regions, like the cerebellum, with greater weight than smaller regions, like the substantia nigra (Figure S1). While the HB-PVS has been shown to be representative of iron accumulation in motor regions, it will not pick up changes in other brain regions.

The HB-PVS (a T2w-MRI derivative measure) is strongly related to iron^6^, and T2w-MRI is an iron-sensitive measure. However, other factors can also affect T2w-MRI signals. We have made efforts to rule out confounding factors like gliosis and edema^25,26,38^; however, other forms of tissue mineralization^67^ or imaging features could impact these measures.

Due to the demographics of the UK, most individuals within this study are of European descent, so it is important to validate these findings in non-European individuals.

## Conclusion

Dietary preferences, especially those related to carbohydrates and alcohol, are associated with measurable differences in iron in motor regions of the brain and with risk for PD. In a typical population, dietary preferences and nutrients associated with decreased iron accumulation in the brain are associated with increased PD risk. These results demonstrate that diet is associated with PD risk and offer a potential mechanistic explanation, suggesting that changes in PD risk may be related to iron levels in motor regions of the brain.

## Data availability

The data used in our analyses was accessed from UK Biobank under accession number 27412. Materials provided by UK Biobank cannot shared according to UK Biobank’s Terms of Use (Section 2.2, https://www.ukbiobank.ac.uk/media/p3zffurf/biobank-mta.pdf).

## Acknowledgements

This work uses data provided by patients and collected by the NHS as part of their care and support. UK Biobank’s research resource is a major contributor in the advancement of modern medicine and treatment, enabling better understanding of the prevention, diagnosis, and treatment of a wide range of serious and life-threatening illnesses – including cancer, heart diseases, and stroke. UK Biobank is generously supported by its founding funders the Wellcome Trust and UK Medical Research Council, as well as the Department of Health, Scottish Government, the Northwest Regional Development Agency, British Heart Foundation, and Cancer Research UK. The organization has over 150 dedicated members of staff, based in multiple locations across the UK.

## Funding

This work was supported by grants R01MH122688 and RF1MH120025, funded by the National Institute for Mental Health (NIMH), and the Lundbeck Foundation fellowship (R335-2019-2318).

## Financial Disclosures and Competing Interests

The authors report no competing interests.

## Author Roles

JA, METB, and RL conceived of the presented idea. JA, WKT, CCF, and RL developed an analysis plan for preference-level, nutrient-level, and factor-level analyses. JA, LS, OA, AD, and RL formulated the analysis plan for the voxel-wise imaging analysis. JA implemented formal analyses and made visualizations. JA and RL wrote the original draft. METB, LS, OA, AD, WKT, and CCF provided comments and revisions. Supervision of this project and administrative oversight were provided by METB, LS, AD, WKT, CCF, and RL. Financial and data resources were provided by CCF, WKT, OA, and AD.

## URLs

UK Biobank: https://www.ukbiobank.ac.uk/

Statsmodels: https://www.statsmodels.org/stable/index.html

FactorAnalyser: https://factor-analyzer.readthedocs.io/en/latest/index.html

## Notes

### Competing Interest Statement

The authors have declared no competing interest.

### Funding Statement

This work was supported by grants R01MH122688 and RF1MH120025 funded by the National Institute for Mental Health (NIMH) in addition to the Lundbeck Foundation fellowship (R335-2019-2318).

### Author Declarations

UK Biobank has approval from the North West Multi-centre Research Ethics Committee (MREC), which covers the UK. It also sought the approval in England and Wales from the Patient Information Advisory Group (PIAG) for gaining access to information that would allow it to invite people to participate. PIAG has since been replaced by the National Information Governance Board for Health & Social Care (NIGB). In Scotland, UK Biobank has approval from the Community Health Index Advisory Group (CHIAG). UK Biobank possesses a Human Tissue Authority (HTA) licence, so a separate HTA licence is not required by researchers who receive samples from the resource, so long as residual samples are destroyed or returned at the end of the research project, and applicants do not transfer the samples to third party premises without the specific approval of UK Biobank. Instead of requiring each applicant to obtain separate ethics approval, UK Biobank has sought generic Research Tissue Bank (RTB) approval, which should cover the large majority of research using the resource. This approach is recommended by the National Research Ethics Service and UK Biobank's governing Research Ethics Committee (REC), which approved the application in 2010. Researchers should check the UK Biobank Access Procedures for more detail.

### Summary of Updates

Updated abstract structure. Moved portions of the analysis to the supplementary materials. Other wording and formatting changes.

