## Supplementary Materials for "Dietary Factors Affect Brain Iron Accumulation and Parkinson’s Disease Risk"

### Supplementary Methods

#### Image Acquisition

Imaging data was accessed from Data-Fields 20216 (T1-MRI) and 20218 (dw-MRI). Images were obtained at three scanning sites in the United Kingdom, on identically configured Siemens Skyra 3T scanners, with 32-channel receive head coils. T1 scans were collected using a 3-dimensional magnetization-prepared rapid gradient-echo (MPRAGE) sequence at 1-mm isotropic resolution. The dw-MRI data were collected using a SE-EPI sequence at 2-mm isotropic resolution using a multishell acquisition. This sequence acquired five b = 0 s/mm^2^ frames and 100 non-collinear gradient directions, with 50 directions at b = 1000 s/mm^2^ and 50 directions at b = 2000 s/mm^2^.

#### Image pre-processing and registration

Preprocessing imaging quality control involves automatic motion detection and expert rating of the imaging quality. The dw-MRI data that passed preprocessing imaging quality control were processed through forward-reverse gradient warping, gradient nonlinearity distortion correction, eddy current correction, and motion correction to reduce the spatial distortion and signal heterogeneities driven by scanner differences. Images were then aligned to a common atlas using rigid-body registration, adjusting the diffusion gradient directions to account for head rotation relative to the atlas as described elsewhere^1^. Fiber orientation density functions were calculated for each voxel, and the derived tensor information, together with T1-MRI information, was fed into multi-channel nonlinear smoothing spline registration, resulting in positional and orientational aligned voxel-wise dw-MRI at 2 mm resolution. Complete details of UK Biobank image acquisition, processing, and quality control can be found in previous work by Alfaro-Almagro and colleagues^1^.

#### Nutrient Data

Nutrient data (Category 100117) consisted of 63 estimates of food nutrient intake based on responses to the Oxford WebQ dietary assessment^2–4^. Items representing a non-typical diet, as indicated by Data-Field 100020, were removed. For participants with more than one estimate of nutritional intake, the mean value of nutritional estimates was used in our analysis. All values were z-score normalized across all subjects.

Statistical analysis used generalized linear models in Python using statsmodels (version 0.14.0), correcting for potential covariates as described in the main text. The age at which the survey was completed was retrieved from Data-Field 105010. P-values were corrected as described in the main text. To calculate robust standard errors across our data, we performed 10,000 bootstrap iterations of our regression analyses^5^ between estimated nutrient intake and the Hemochromatosis Brain PolyVoxel Score (HB-PVS), and estimated nutrient intake and the odds ratio for PD.

To understand potential joint effects of nutrient intake, central iron levels, and PD risk we performed instrumental variables regressions where we plotted effect sizes and identified correlations that would indicate patterns of related associations as described in the main text. This analysis used regression coefficients and standard errors from preceding analyses (described above) capturing the relationship between a) estimated nutrient intake and central iron accumulation, and b) estimated nutrient intake and PD risk.

### Supplementary Results

#### Dietary/Lifestyle Preferences and Brain Iron

To understand how food and lifestyle preferences may be related to brain iron accumulation, we performed a fit of a series of linear regression models associating each diet/lifestyle preference with the HB-PVS. The strongest positive associations were between wine and the HB-PVS (red wine: t=6.72, p_FDR_<1e-8; white wine: t=5.48, p_FDR_<1e-5). There were additional FDR significant positive associations between the HB-PVS and other alcoholic beverages, including spirits (t=3.56, p_FDR_=0.004) and whiskey (t=2.83, p_FDR_=0.022). Each of these indicates that a greater preference for alcohol is related to greater iron accumulation in motor regions captured by the HB-PVS.

There were also Bonferroni significant positive associations between the HB-PVS and preferences for red meats as a collective category (t=4.46, p_FDR_<0.001) and individual red meats like lamb (t=4.30, p_FDR_<0.001) and beef steak (t=4.17, p_FDR_<0.001) among others. Additional positive associations between meats and the HB-PVS were (in order of descending significance) grilled meat, sausage meat, pork chop, bacon, roast chicken, ham, non-prawn shellfish, prawns, burger meat, salami, and chicken (Figure S2 and Table S5). Once again, preferences for these foods indicate greater brain iron accumulation.

There were significant negative associations between the HB-PVS and food made from cereal grains like corn flakes (t=-4.61, p_FDR_<0.001), porridge (t=-3.74, p_FDR_=0.004), and cereal granola bars (t=-3.36, p_FDR_=0.007). There were also significant negative associations between the hemochromatosis brain and sweets, including biscuits (t=-3.64, p_FDR_=0.004), cake (t=-3.56, p_FDR_=0.004), milk chocolate (t=-3.36, p_FDR_=0.007), sweet coffee house drinks (t=-3.08, p_FDR_=0.014), and cake icing (t=-2.64, p_FDR_=0.031).

There is also an FDR negative association between the HB-PVS and preferences for forms of exercise, including bicycling (t=-3.19, p_FDR_=0.010), exercising alone (t=-3.03, p_FDR_=0.015), working up a sweat (t=-2.78, p_FDR_=0.024), and taking the stairs (t=-2.57, p_FDR_=0.035). These results indicate that a preference for certain breakfast cereals, high-sugar foods/drinks, and physical exercise is associated with lower levels of iron in motor circuits of the brain.

A graphical representation of these results can be found in Figure S2, and a full numeric summary of all the relationships between preferences and the HB-PVS can be found in Table S5.

#### Dietary/Lifestyle Preferences and Parkinson’s Disease Risk

We fit a series of linear regression models to understand how preferences might be associated with PD risk. Four main categories of preferences were associated with PD risk: exercise, fruits/vegetables, alcoholic beverages, and sweets.

We found that increased PD risk was associated with a preference for less exercise including taking the stairs (t=-10.87, p_FDR_<1e-24), working up a sweat (t=-7.67, p_FDR_<1e-12), and exercising alone (t=-7.21, p_FDR_<1e-10). We suspect that as movement deficits are a core feature of the disorder these results may be an example of reverse causation where those living with PD develop a preference for not exercising leading to the significant results we observe.

We also identified that lower PD risk was associated with a preference for produce including a collective preference for vegetables (t=-6.24, p_FDR_<1e-7) and several individual vegetables including salad leaves (t=-8.27, p_FDR_<1e-13), onions (t=-5.65, p_FDR_<1e-6), lentil beans (t=-5.58, p_FDR_<1e-5), as well as (in descending order of significance) cucumber, cabbage, cauliflower, white turnip, spinach, broccoli, broad beans, fresh tomatoes, pears, asparagus, beetroot, avocados, plums, grapefruit, and garlic (Figure S3 and Table S6).

We identified a lower PD risk was associated with a preference for alcohol including spirits (t=-6.15, p_FDR_<10e-7), red wine (t=-5.80, p_FDR_<10e-6), whiskey (t=-3.50, p_FDR_=0.002), bitter ale (t=-3.33, p_FDR_=0.004), going to the pub (t=-2.94, p_FDR_=0.012), and white wine (t=-2.66, p_FDR_=0.024).

We identified higher PD risk was associated with a preference for sweet foods as a category (t=4.52, p_FDR_<1e-4) and individual sweet foods including sweet coffee house drinks (t=5.15, p_FDR_<1e-5), cake icing (t=4.90, p_FDR_<1e-4), cheesecake (t=4.81, p_FDR_<1e-4), as well as (in order of descending significance) biscuits, regular non-diet fizzy drinks, ice cream, milk chocolate, cake, marzipan, jam, and apple juice (Figure S3, Table S6).

A graphical representation of these results can be found in Figure S3. A full numeric summary of all the relationships between preferences and the HB-PVS can be found in Table S6.

#### Non-Dietary Preferences, Brain Iron and Parkinson's Disease Instrumental Variable Regression

Across dietary and non-dietary preferences, we do not find a significant correlational relationship (r=-0.129, p=0.125) (Figure S8). We believe the difference between the analysis based solely on dietary preferences and the one including non-dietary preferences questions are related strongly to exercise which we believe may be related to PD risk through reverse causation (PD patients having a lower preference to exercise because of movement deficits) - hence our treatment of dietary and non-dietary preferences separately.

We see a broad pattern emerging within dietary measures where preferences toward sweet foods are related to lower HB-PVS (lower iron levels in motor circuits) and higher PD risk, while alcohol-related preferences are associated both with higher HB-PVS (higher iron levels in motor circuits) and lower PD risk (Figure 1A).

#### Estimated Nutrient Intake and Brain Iron

To understand how estimated nutrient intake is associated with the pattern of iron accumulation captured by the HB-PVS, we performed linear regressions in our imaging sample (N=20,477). We identified several FDR-corrected significant negative associations between the HB-PVS and starch (t=-3.21, p_FDR_=0.018), carbohydrates (t=-3.27, p_FDR_=0.013), riboflavin (t=-3.27, p_FDR_=0.013), vegetable protein (t=-3.31, p_FDR_=0.013), lactose (t=-3.36, p_FDR_=0.013), and maltose (t=-3.74, p_FDR_=0.012) (Figure S4A, Table S3) - indicating higher consumption of these nutrients is linked with lower levels of brain iron in motor regions captured by the HB-PVS.

#### Estimated Nutrient Intake and Parkinson’s Disease Risk

To examine the relationship between estimated nutrient intake and risk of developing PD we performed a linear regression in our non-imaging sample (N_Case_=979 and N_Control_=149,624). We found estimated intake of energy beverages (t=-2.87, p_FDR_=0.037) and alcohol (t=-4.56, p_FDR_<0.001) were negatively associated with PD risk. Estimated higher consumption of free sugar (t=2.73, p_FDR_=0.044), fructose (t=2.82, p_FDR_=0.038), sucrose (t=3.06, p_FDR_=0.023), non-milk extrinsic sugars (t=3.19, p_FDR_=0.018), glucose (t=3.26, p_FDR_=0.017), total sugar (t=3.45, p_FDR_=0.012), and carbohydrates (t=3.46, p_FDR_=0.012) were all positively associated with PD risk (Figure S4B, Table S4).

#### Estimated Nutrient Intake, Brain Iron, and Parkinson's Disease Instrumental Variable Regression

To understand the joint effect of nutrients on brain iron accumulation, as captured by the HB-PVS, and PD risk we performed instrumental variable regression on our previous analyses generated in non-overlapping samples (Table 1). We identified a significant negative association (r=-0.421, p=0.0006) between a) nutrient intake and brain iron accumulation and b) nutrient intake and PD risk (Figure S4C). This suggests that nutrient-mediated increases in brain iron are linked with lower PD risk. Many sugar-related items simultaneously display a pattern of increased PD risk and decreased brain iron accumulation (as captured by the HB-PVS) (upper left of Figure S4C).

#### Incidental Free Water Findings

The primary purpose of our free water analysis was to interrogate the degree to which our T2w-MRI associations with factors 2 (sweets) and 7 (alcohol) could be driven by vascular pathologies. Preference factors for sweets and alcohol were associated with basal ganglia and cerebellar intensities for T2w-MRI imaging but not free water estimates (compare the upper and lower panels of Figure 2). From these findings, contextualized with previous evidence of association with iron regulatory phenotypes^6,7^, we conclude that observed alcohol and sweet T2w-MRI effects are more likely related to iron processes than being confounded by vascular disruptions.

Separate from our findings in T2w-MRI, we identify several free water hyper- and hypointensities in ventricular regions in the brain related to factor 2 (sweets) and factor 7 (alcohol), respectively. We approach the interpretation of these results with caution^8^; however, differences may be related to the impacts that diet may have on CSF composition and permeability. Differences in diet are related to differences in CSF secretion^9^ and changes in the levels of various biomarkers in the CSF^10^, including those related to Alzheimer’s disease^11^. Alcohol specifically can also alter CSF composition biomarkers^12^, including those related to Alzheimer’s Disease^13,14^, and can increase blood-brain barrier permeability^15^. Further research should delve deeper into studying CSF composition along these preference factors and separate these findings from potential differences in surrounding tissue structure^8^.

### Supplementary Discussion

#### Exercise

We found that preferences for increased exercise were associated with reduced PD risk and reduced brain iron as measured with the HB-PVS. This result is discordant with the trends observed for estimated nutrient intake and dietary preferences, where items that were linked to increases in brain iron were related to decreases in PD. We suspect that this discordant result may be spurious due to the negative exercise and PD association being likely driven by reverse causation - e.g. the core motor deficits of the disease make physical activity less desirable - hence the distinction between dietary and non-dietary measures in our instrumental variable regression. Nevertheless, the association between exercise and brain iron levels in motor regions appears to be consistent with other research indicating exercise can modulate iron stores in the body by both changing internal rates of hemolysis^16^ as well as modulating hepcidin^17^. Although we believe reverse causation explains the discordant result of exercise being associated with reductions in both brain iron and PD risk, it is possible that our findings are not a result of reverse causation. Indeed, moderate to high levels of activity were found to be associated with a lower risk for developing PD later in life, and individuals with PD who report higher physical activity were found to have slower symptom progression and better quality of life^18,19^. Population analyses in UK Biobank have also found that sedentary behavior is associated with an increased incidence of dementia^20^. However, in these studies, it is difficult to account for survivorship bias. Animal models of Alzheimer's disease have found that treadmill exercise helps to alleviate disease-related iron dysregulation as well as disease-induced cognitive decline and neuronal death^21^. More research is needed to understand the direction of causation of effects and the mechanisms behind the apparent benefit of physical exercise for PD patients.

#### Additional Factors that Influence PD Risk

Analysis showed that preferences for vegetables and fruits were associated with decreased risk of PD, and no significant association with the HB-PVS. These results are consistent with the findings from previous studies that found high fruit and vegetable intake is related to lower PD risk^22,23^. High consumption of fruits and vegetables could explain a portion of the PD-protective effects seen in the Mediterranean diet^24–26^. We believe that our results are consistent with a beneficial effect of vegetable and fruit intake on PD risk that does not converge on altered central iron levels and may instead result from bioactive compounds in vegetables that may reduce PD risk in an iron-agnostic way. Furthermore, a diet rich in fruits and vegetables fosters the growth of short-chain fatty acid-producing bacteria in the gut microbiota, which can reduce gut permeability, lower inflammation, and decrease PD risk^27^.

#### Additional Factors that Influence the HB-PVS

We also identified some factors that affected the HB-PVS but not risk of PD. We found that preferences related to meat (preferences for red meat, lamb, beef steak, grilled meat, etc., and Factor 1), especially red meat, were associated with increased measures of the HB-PVS - i.e. greater iron levels in motor regions. This is consistent with previous findings that vegetarians have markers indicative of depleted iron stores and a higher proportion of iron deficiency anemia compared to non-vegitarians^28^. Red meat, especially beef, contains high concentrations of highly bioavailable heme iron and animal proteins, and as such, red meat consumption is not recommended for those at risk of iron overload^29,30^. The relationship between red meat consumption and neurodegeneration and PD is still unclear. Our research identified a significant association between a preference for beef steak and an increased brain iron in motor regions as well as a decreased risk of PD; however, recent systematic reviews have found no association between red meat consumption and cognitive impairment, Alzheimer’s disease, dementia, or PD^31,32^. The actual iron absorbed from meats will vary based on the cut and cooking method^33^ as well as the presence or absence of iron absorption inhibitors or promoters in the meal^34^. Future research should overcome this by carefully assessing different methods of quantifying red meat consumption or working to understand the effects of diet at a nutrient level.

Our analyses found that preferences related to cereal grains were associated with reduced brain iron as captured by the HB-PVS. This result is surprising given that cereal is a common vehicle for added iron fortification. As discussed above, literature has demonstrated that the ingestion of sugars increases hepcidin production and decreases iron absorption^35^, which may explain our seemingly contradictory results. With cereal grains containing 50-80% carbohydrate^36^, it is possible that high sugar consumption with cereal is leading to lower iron absorption, which overcomes the effects of iron fortification. Additionally, cereals and dairy products, usually consumed alongside cereals, are high in iron-absorption inhibitors like phytic acid and calcium, which reduce iron bioavailability by chelating and cloistering iron in the digestive tract^37^. Our current interpretation is that lower brain iron levels observed in individuals preferring cereal grains are due to iron-absorption inhibitors in these meals and that the association we observe occurs despite cereal iron fortification, not because of it.

### Figure S1: Hemochromatosis Brain PolyVoxel Score Region Weighting

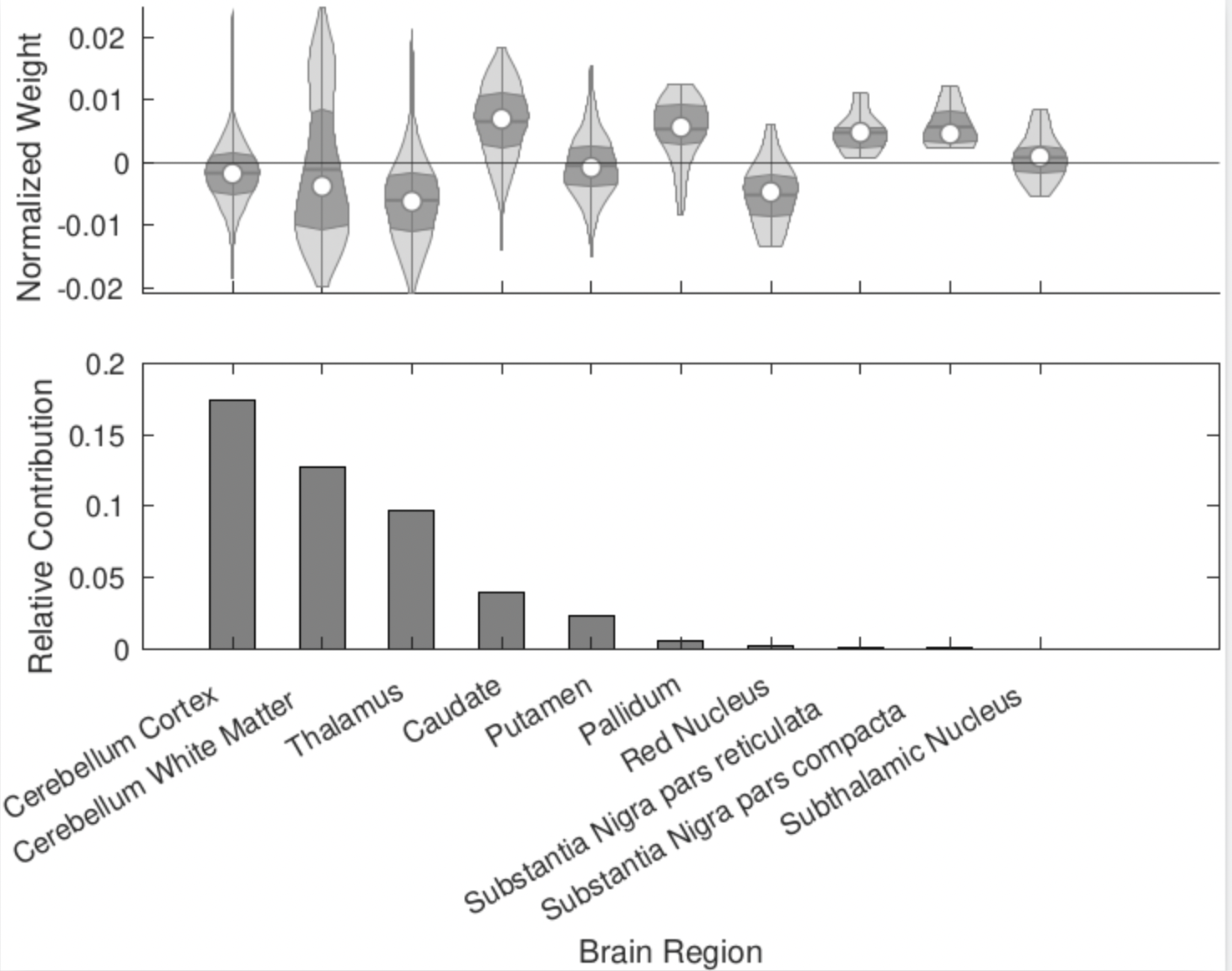

**Figure S1**: Hemochromatosis Brain PolyVoxel Score (HB-PVS) weight breakdown by each region of interest (ROI). The top plot represents the posterior weights by brain region. Weights are normalized to be of unit length. The bottom plot shows the relative contribution of each region to the HB-PVS as the sum of squares within each brain region for posterior weights. This plot is adapted from the work done by Loughnan et. al. 2024^7^.

### Figure S2: Associations Between Food Preferences and Hemochromatosis Brain PVS

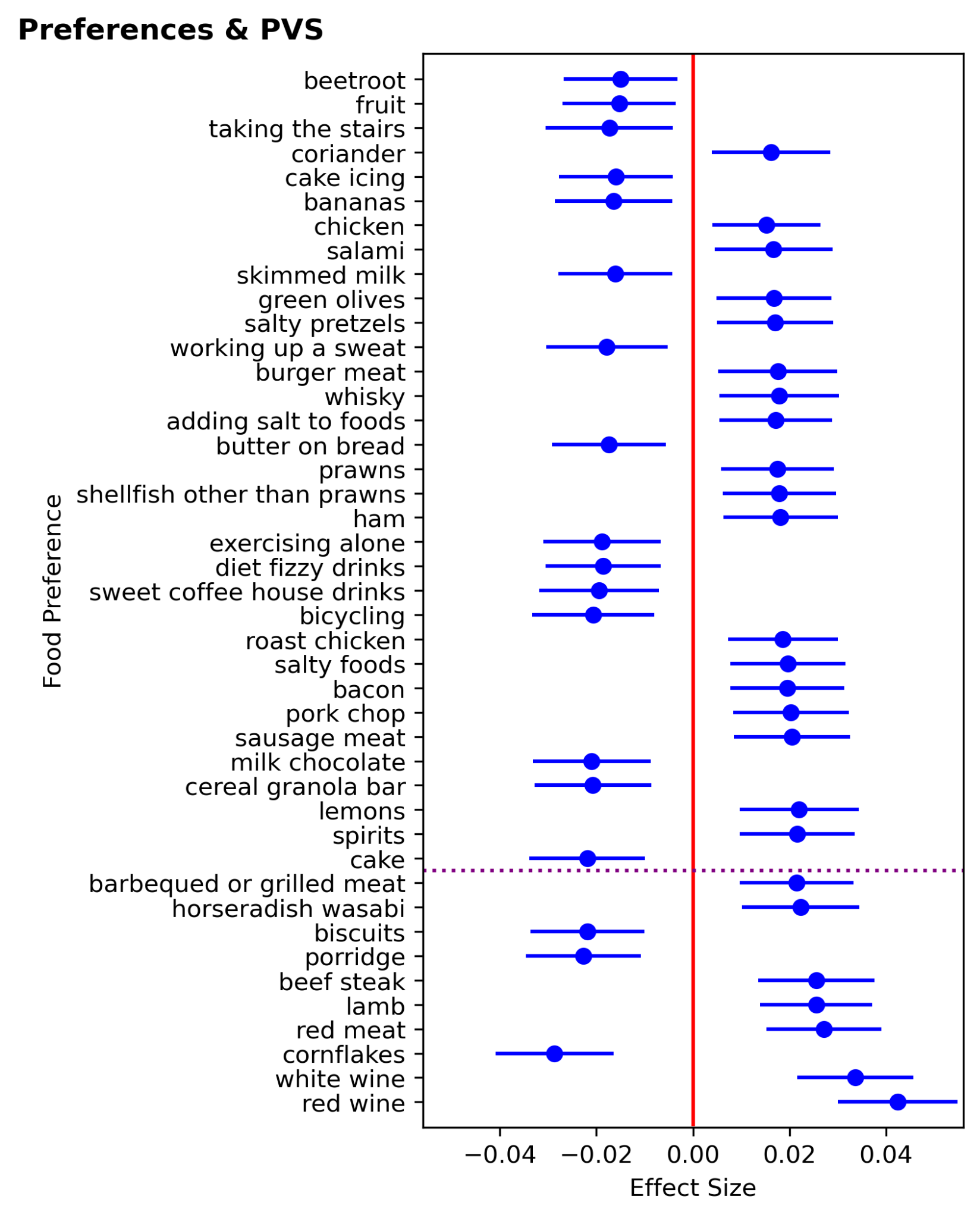

**Figure S2**: Forrest plot showing the effect size and 95% confidence intervals of FDR significant preferences and the iron-sensitive measure, the HB-PVS. Results below the purple line are additionally Bonferroni Significant. These results are also shown in tandem with their associations with PD risk in Figure 2A. A full numeric summary of all the relationships between preferences and the HB-PVS can be found in Table S5.

### Figure S3: Associations Between Food Preferences and PD Risk

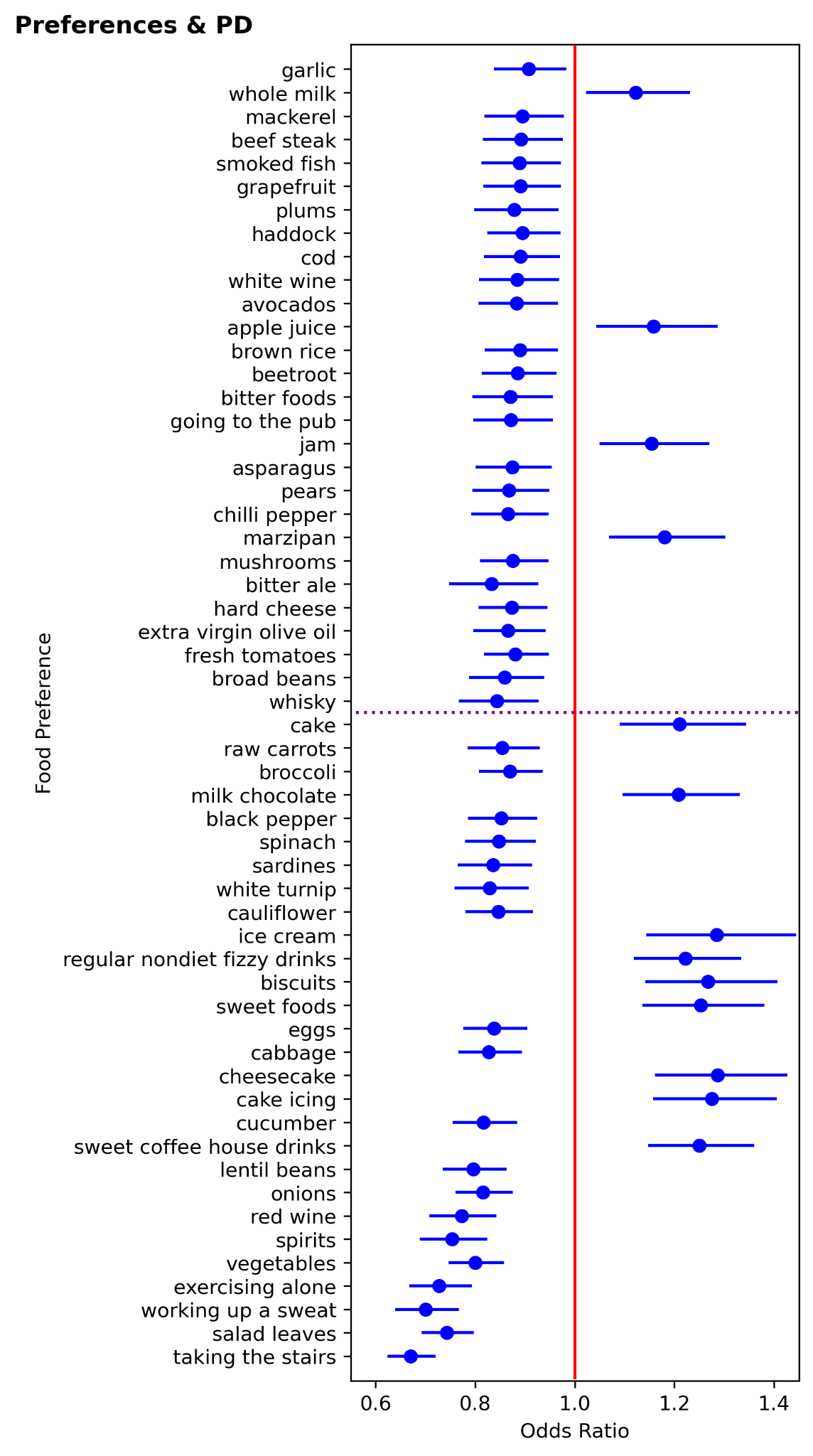

Figure S3: Forrest plot showing the effect size and 95% confidence intervals of FDR significant preferences and odds ratio of PD. Results below the purple line are additionally Bonferroni Significant. These results are also shown in tandem with their associations with the HB-PVS in Figure 2A. A full numeric summary of all the relationships between preferences and the HB-PVS can be found in Table S5.

### Figure S4: Nutrient Analysis

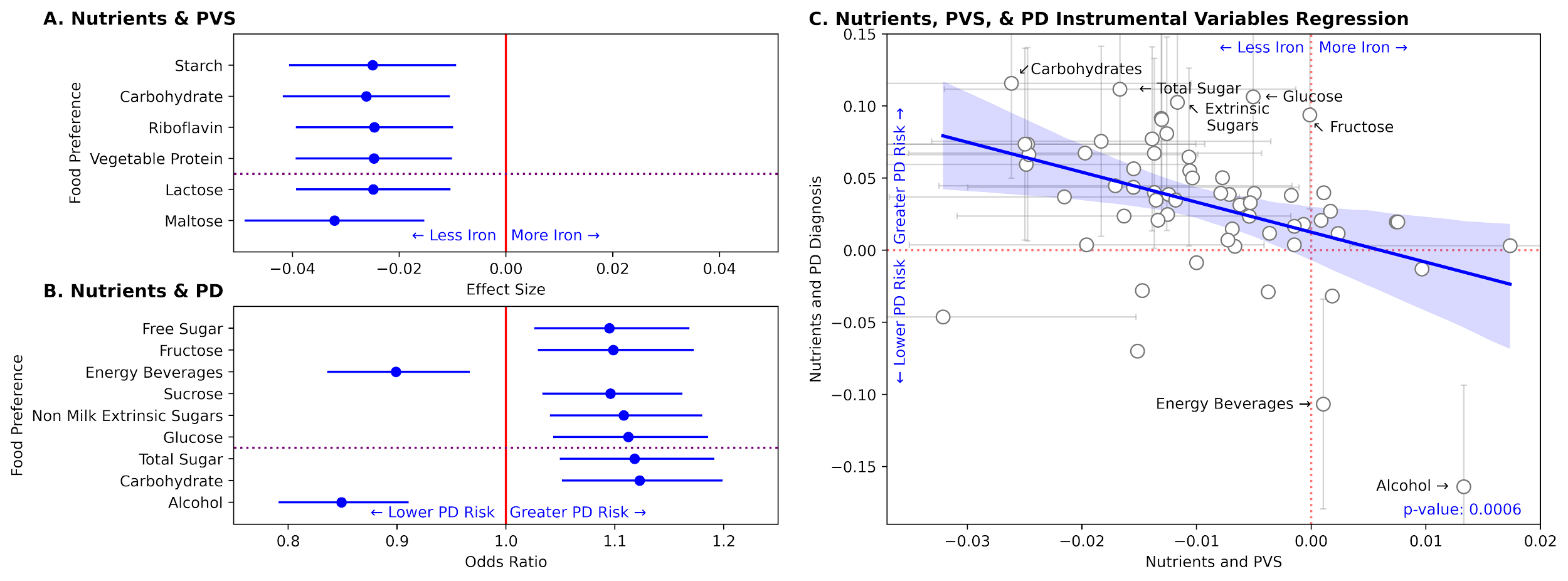

**Figure S4**: (A) Forest plot showing the FDR significant associations between estimated nutrient intake and the HB-PVS. Results below the dotted line are additionally Bonferroni Significant. A full numeric summary of all the results analyzed can be found in Table S3. (B) Forest plot showing the FDR significant associations between estimated nutrient intake and odds ratio for developing PD. Results below the dotted line are additionally Bonferroni Significant. A full numeric summary of all the results analyzed can be found in Table S4. (C) Scatter plot of the instrumental variable regression between a) estimated nutrient intake and the HB-PVS and b) estimated nutrient intake and PD risk. The top seven nutrients furthest from the origin are annotated. Instrumental variable regression revealed a significant negative relationship, highlighting that nutrients increasing brain iron levels were related to decreased PD risk. Error bars are included for results that are significant on a given axis.

### Figure S5: Dendrogram of Preferences

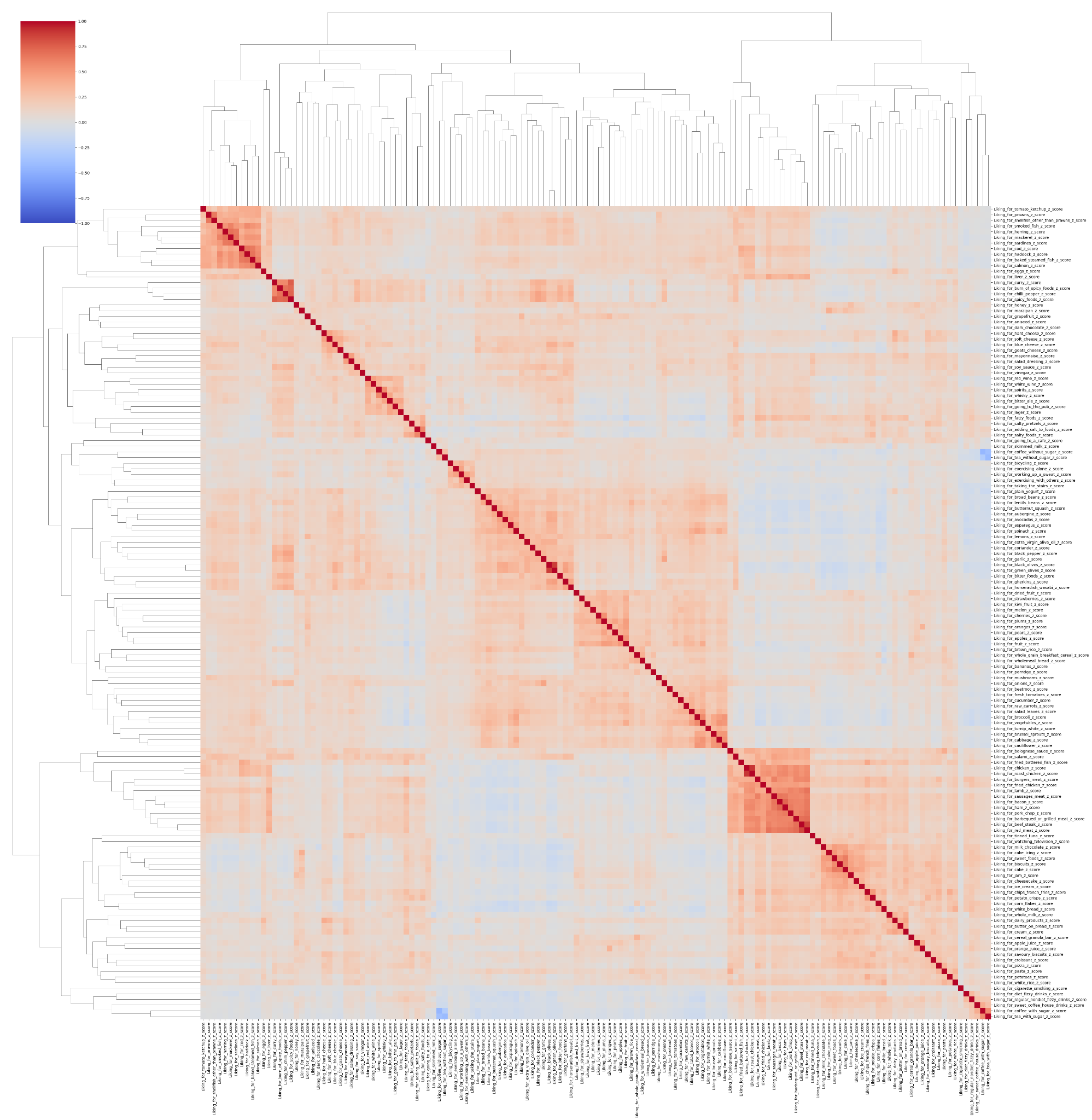
**Figure S5:** This is a dendrogram showing a hierarchical clustering of the preferences based on how similar they are. The color gradients represent the correlation.

### Figure S6: Scree Plot for the Factor Analysis of Preference Data

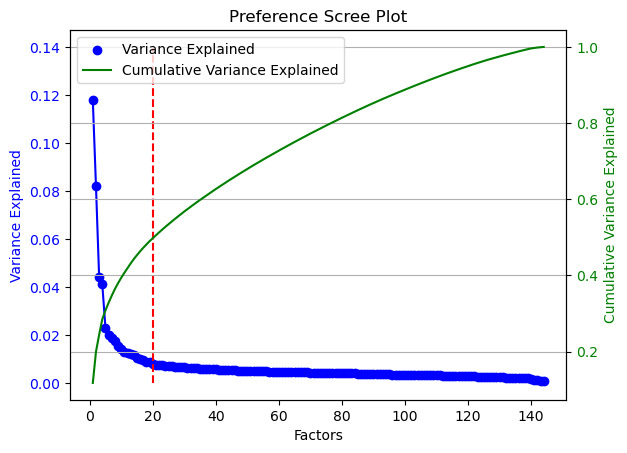

**Figure S6**: Scree plot showing the variance explained by factor and the cumulative variance explained. The red, dotted line indicates the chosen number of factors for our analysis.

### Figure S7: Heatmap of Preference Factor Loadings

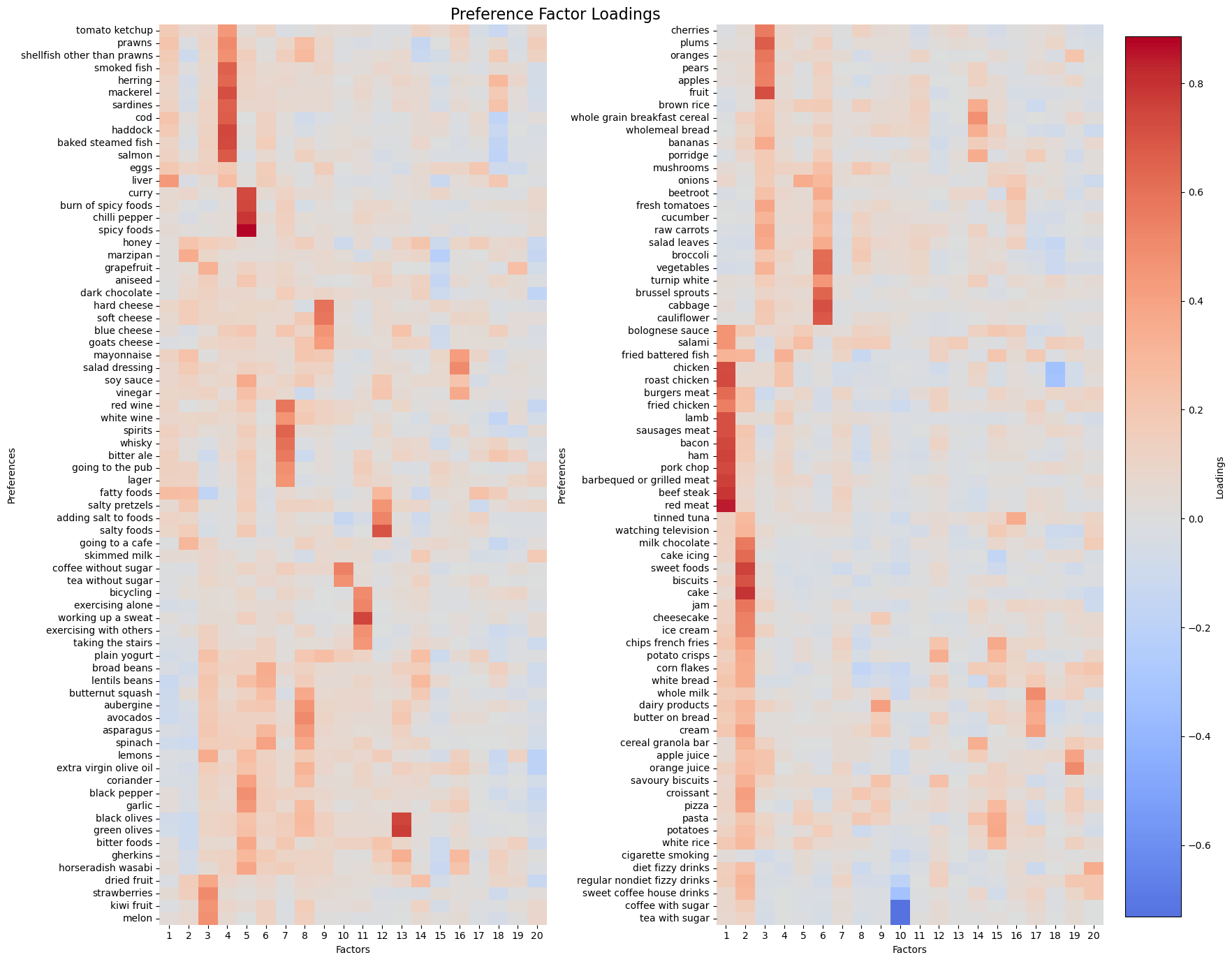

**Figure S7:** Heatmap indicating the positive and negative factor loadings associated with each factor.

### Figure S8: Preferences, PD, & HB-PVS Instrumental Variables Regression

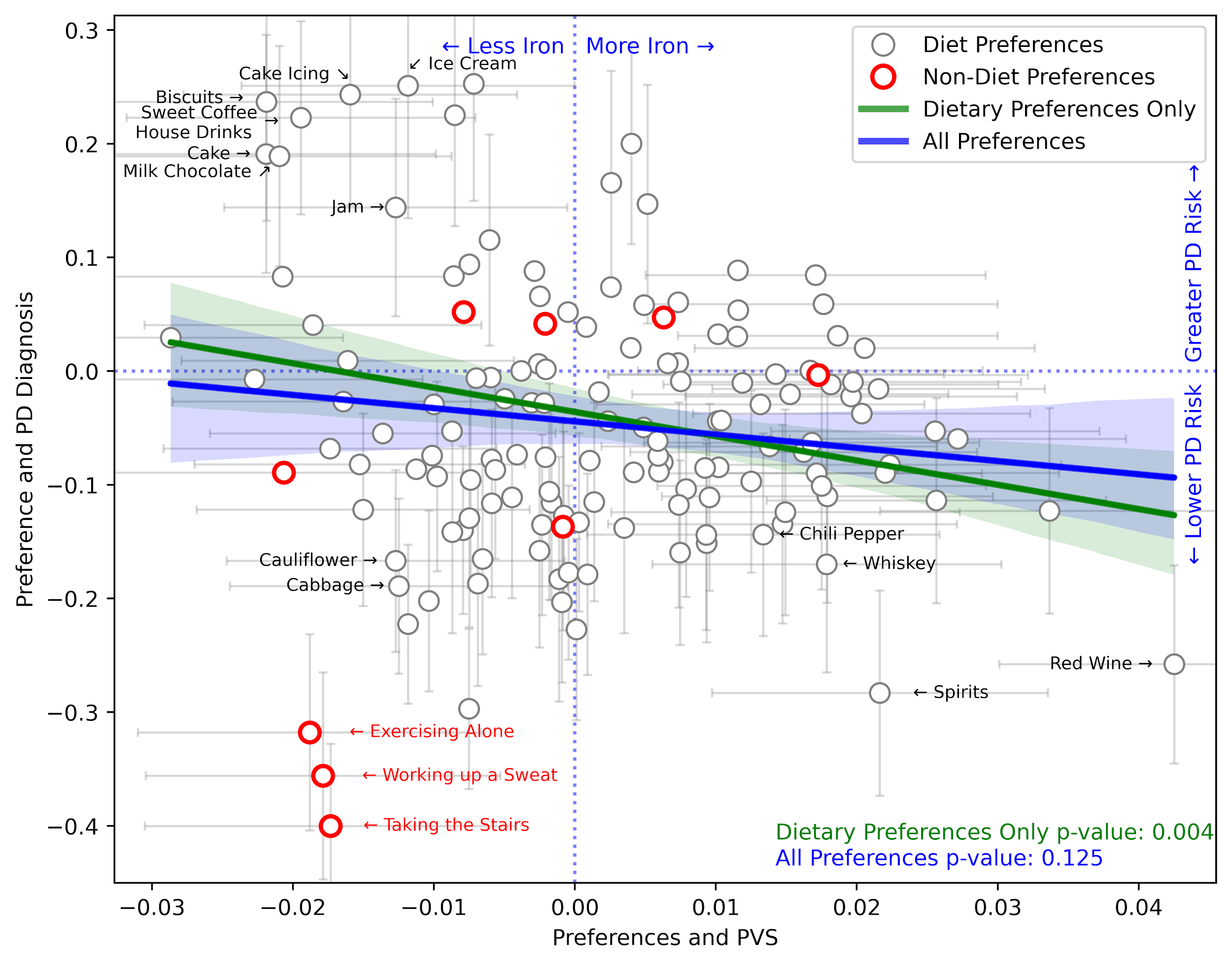

**Figure S8:** Scatter plot of the instrumental variable regression between preferences and the HB-PVS and preferences and the odds ratio of developing PD. Error bars are shown for factors that are significant along a given axis. Of those factors, the sixteen preference factors furthest from the origin are annotated. Regression lines show correlations between the effects of a preference on PD and the HB-PVS. Non-dietary factors (marked in red) include preferences for taking the stairs, working up a sweat, exercising alone, bicycling, going to a cafe, watching television, cigarette smoking, exercising with others, and going to the pub.

### Figure S9: Unmasked Dietary Factor Association Maps**Figure S9:** Voxel-wise association map of T2-w signal intensity for i) factor 2 (sweets), ii) and factor 7 (alcohol). Association maps display beta effects; orange regions represent increased T2-weighted intensities for individuals with a higher scoring on the spectrum of a given dietary factor and blue regions represent decreased T2-weighted intensities.

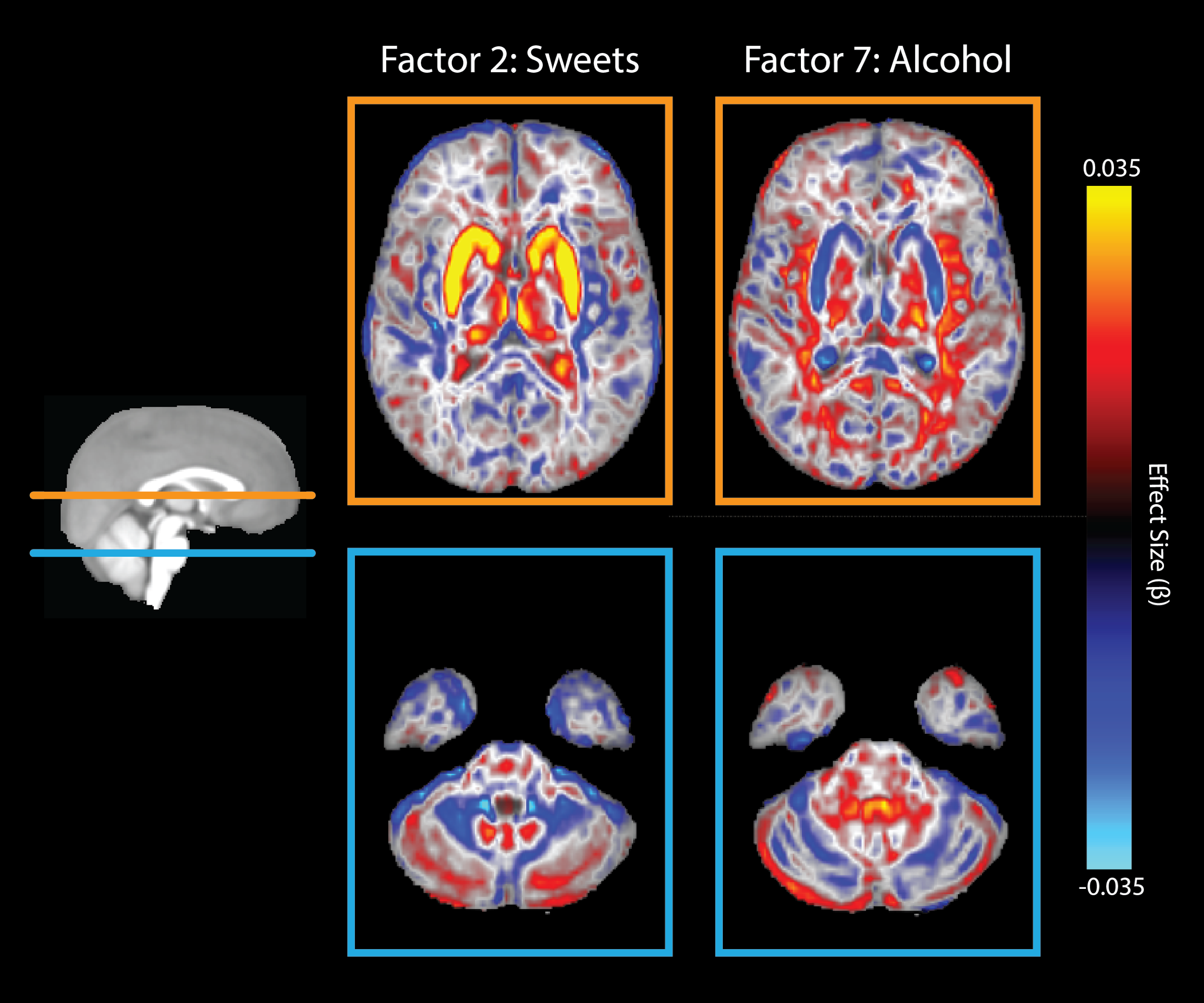
